# Characterizing Affective Variability in Bipolar Disorder and Borderline Personality Disorder, and the Effects of Lithium, Using a Generative Model of Affect

**DOI:** 10.1101/2022.02.18.22271166

**Authors:** Erdem Pulcu, Kate E.A. Saunders, Catherine J. Harmer, Paul J. Harrison, Guy M. Goodwin, John R. Geddes, Michael Browning

## Abstract

The affective variability of Bipolar Disorder (BD) is thought to qualitatively differ from that of Borderline Personality Disorder (BPD), with changes in affect persisting for longer in BD. However, quantitative studies have not been able to confirm this distinction. It has therefore not been possible to accurately quantify how treatments like lithium influence affective variability in BD. We assessed the affective variability associated with BD and BPD as well as the effect of lithium using a novel computational model that defines two subtypes of variability: affective changes that persist (volatility) and changes that do not (noise). We hypothesized that affective volatility would be raised in the BD group, noise would be raised in the BPD group and that lithium would impact affective volatility. Daily affect ratings were prospectively collected for up to 3 years from patients with BD, BPD and non-clinical controls. In a separate experimental-medicine study, patients with BD were randomized to receive lithium or placebo, with affect ratings collected from week -2 to +4. We found a diagnostically specific pattern of affective variability. Affective volatility was raised in patients with BD whereas affective noise was raised in patients with BPD. Rather than suppressing affective variability, lithium increased the volatility of positive affect in both studies. These results provide a quantitative measure of the affective variability associated with BD and BPD. They suggest a novel mechanism of action for lithium, whereby periods of persistently low or high affect are avoided by increasing the volatility of affective responses.

## Introduction

Excessive affective variability, sometimes called affective instability, characterizes psychiatric diagnoses such as bipolar disorder and borderline personality disorder (1–4), and is associated with adverse outcomes across diagnoses (5, 6). It has been suggested that affective instability may be an important treatment target across a range of psychiatric presentations (3, 7, 8).

However, different types of affective variability are thought to exist; when asked to retrospectively describe affective variability, patients with bipolar disorder report longer periods of raised or lowered affect whereas patients with borderline personality disorder report a higher frequency variation of affect (4). Consistent with this difference, mood stabilizing medications like lithium, which reduce the occurrence of mania and depression (i.e. particularly prolonged periods of extreme affect) in bipolar disorder (9), have not been found to be effective in patients diagnosed with borderline personality disorder (10).

Affective variability may be directly estimated from prospectively collected affect ratings, with a variety of different metrics of variability described (11–14). However, the different measures of variability tend to be highly correlated with each other (14) and to date have not been able to capture the qualitative differences in affective variability described for bipolar and borderline personality disorders. For example, the same measures of affective variability that are raised in borderline personality disorder (11, 15–17) are also raised in bipolar disorder (11), post-traumatic stress disorder and bulimia nervosa (16). Existing measures of variability of affect ratings therefore lack diagnostic specificity and cannot account for differences in treatment response between diagnoses.

An alternative approach to conceptualizing and measuring the variability of an outcome is to construct a generative model of how that outcome is produced and then to invert the model using Bayes rule (18, 19). A generative model formally describes the assumed causal processes that produce an outcome (Figure 1), inversion of the model creates a “Bayesian filter” (18–21) which allows one to start with the observations and then to estimate distinct, model-defined, causes of variability within a single, overarching framework.

**Figure 1.**
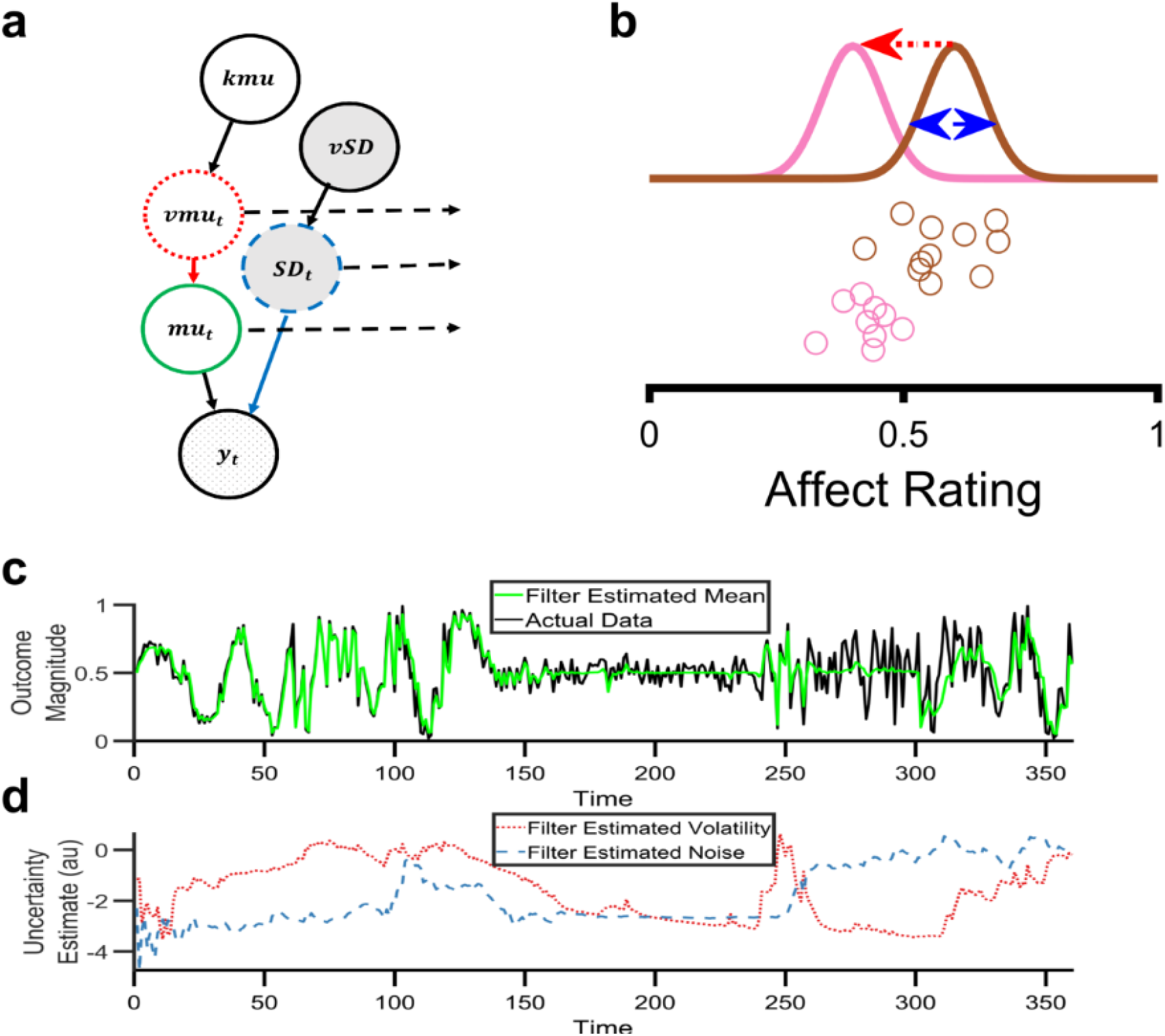
Figure 1: A Bayesian filter to estimate types of affective variability. **a**: A graphical illustration of the generative model which describes how affect ratings (represented by *y*_*t*_) are produced at each time point. The hypothesized causal processes leading to the production of the ratings is controlled by the nodes, *mu*_t_, *SD*_*t*_, *vmu*_*t*_, *kmu* and *vSD*, which are described in the main text. **b**: An illustration of the types of variability in the generative model. Circles represent individual affect ratings, sequentially generated from top to bottom. The color of the circle indicates the distribution from which it was drawn. One type of variability, volatility (*vmu*_*t*_, red arrow), arises from a shift in the distribution (from brown to pink), leading to a change in all subsequent ratings. A second type of variability, noise (*SD*_*t*_, blue arrow), arises from the sampling of the ratings from the distributions and leads to independent changes in each rating. **c**: Behaviour of the Bayesian filter using synthetic data. The black line illustrates a timeseries of synthetic data drawn from the range 0-1. The data contains periods where volatility is high (time 1-120 and 301-360) and others where it is low (time 121-300). Similarly, it contains periods in which noise is high (time points 61-120 and 241-360) and low (time 1-60 and 121-240). The green line illustrates the Bayesian filter’s belief about the mean of the generative process, *mu*_*t*_, at each timepoint. As can be seen, the filter changes its estimate of the mean when it thinks variability in the data is caused by volatility (e.g. time 1-60) and does not alter its estimate of the mean when it thinks variability is caused by noise (e.g. time 260-300). It is able to adapt to changes in the level of volatility and noise, although occasionally misattributes the cause (e.g. when the noise increases at time 240, the filter initially believes this is caused by an increase in volatility before correctly attributing it to noise by time 260). **d:** The filter’s estimate of volatility (red line) and noise (blue line) from the same synthetic data as **c**. Panels **c** and **d** are adapted from(19).

In this paper we inverted a simple generative model of affect (Figure 1) to estimate two different causes of affective variability; volatility which leads to persistent change in affect and noise which leads to transient change. We applied this approach to prospectively collected affect ratings of patients with bipolar and borderline personality disorders as well as control subjects to assess whether it was able to capture the qualitative differences in affective variability between these diagnostic groups. We then used the model to characterize the causal effects of lithium on affective variability in an experimental medicine study of patients with bipolar disorder. We hypothesized that bipolar disorder would be associated with increased affective volatility, borderline personality disorder with increased affective noise and that lithium would impact affective volatility.

## Results

The generative model of affect and associated Bayesian Filter are summarised in Figure 1. A detailed description and assessment of the performance of the filter is provided in the methods and supplementary information. The key feature of the filter is that it estimates two forms of variability; that caused by affective volatility (i.e. changes of affect that persist over time) and that caused by affective noise (i.e. changes in affect that are transient). We first used the filter to characterise prospectively collected, daily, positive and negative affect ratings (11) from a cohort study of patients with diagnoses of bipolar disorder (n=53), borderline personality disorder (n=33) and non-clinical controls (n=53), see Table 1.

**Table 1.** Demographic details of patients included in the two studies.

| *Cohort Study* |  |  |  |  |
| --- | --- | --- | --- | --- |
|  | **Bipolar Disorder** | **Borderline Personality Disorder** | **Control** | **p value for group difference** |
| **n** | 51 | 33 | 51 |  |
| **Sex (F/M)<sup>a</sup>** | 32/19 | 30/2 | 32/18 | 0.004* |
| **Age (mean(SD))** | 39.47<br>(13.03) | 33.72 (10.42) | 38.18<br>(12.98) | 0.12 |
| **Educational Achievement (mean(SD))<sup>b</sup>** | 4.81<br>(0.96) | 4.1 (0.88) | 5.1<br>(1.02) | <0.001* |
| **Previous Hospitalisation (Y/N)** | 21/28 | 14/17 | 0/48 |  |
| **Subtype of Bipolar Disorder (I/II)** | 30/17 | N/A | N/A |  |
| **On Any Psychoactive Medication (Y/N)** | 48/3 | 24/8 | 1/50 |  |
| **On Lithium (Y/N)** | 22/29 | 0/32 | 0/51 |  |
| **On Anticonvulsant (Y/N)** | 19/32 | 1/31 | 0/51 |  |
| **On Antipsychotic (Y/N)** | 34/17 | 6/26 | 0/51 |  |
| **On Antidepressant (Y/N)** | 16/34 | 23/9 | 1/50<sup>d</sup> |  |
| **On Anxiolytic (Y/N)** | 2/48 | 8/24 | 0/51 |  |
| **On Hypnotic (Y/N)** | 4/46 | 2/29 | 0/51 |  |
| **Current or Previous Psychotherapy (Y/N)<sup>e</sup>** | 42/7 | 31/1 | 11/33 |  |
| **Current or Previous Recreational Drug Use (Y/N)** | 29/18 | 21/10 | 12/38 |  |
| **Experimental Medicine Study** |  |  |  |  |
|  | **Lithium Group** | **Placebo Group** |  |  |
| **N** | 19 | 16 |  |  |
| **Sex (F/M)** | 8/11 | 7/9 |  |  |
| **Age in years (mean(SD))** | 28.84 (9.81) | 35.14 (13.79) |  |  |
| **Diagnosis (BP I/BP II/BP NoS)** | 3/16/0 | 3/11/2 |  |  |

<sup>a</sup> Borderline group differed significantly from both other groups. Bipolar and control groups did not differ.
<sup>a</sup> Demographic data was missing from some participants: one patient in the borderline group did not provide any demographic data, data on educational and employment level was missing from 3 patients in the bipolar group, and 2 patients in the borderline group. Binary outcomes (e.g. sex, medications taken etc) are reported for all participants who provided the relevant data. <sup>b</sup> Educational achievement was rated on a six-point scale from primary school to post graduate level. SD= Standard deviation, BP I/II/NoS = Bipolar disorder type 1, type 2, not otherwise specified. <sup>d</sup> One participant in the control group was taking low dose amitriptyline for pain. <sup>e</sup> Including any form of therapy or counselling.

### Distinct Types of Affective Variability in Bipolar and Borderline Personality Disorders

When considering the standard summary statistics (14) of the affective ratings from the three cohorts, the average ratings of positive affect did not differ between groups [*F*(2,132) = 1.52, *p* = 0.26], although negative ratings did differ [*F*(2,132) = 32.26, *p* < 0.001] with patients in the borderline personality group endorsing higher mean ratings than both of the other groups [both *p*_*bonf*_*’*s < 0.006], and patients in the bipolar group providing higher ratings than the control group [*p*_*bonf*_ < 0.001]. An identical ordering of the groups was apparent for both positive [*F*(2,122) = 11.6, *p* < 0.001] and negative [*F*(2,122) = 38.2, *p* < 0.001] affective variability, as estimated using the standard deviation of the ratings (Figure 2, panels a-d) and, as previously reported, other measures of variability including the root mean square of successive differences, the entropy and the Teager-Kaiser Energy Operator (11). Thus, while the magnitude of the variability metrics differed between groups, there was no specific association between qualitative types of variability and diagnosis, with all measures being higher in the borderline personality group than the bipolar group.

**Figure 2.**
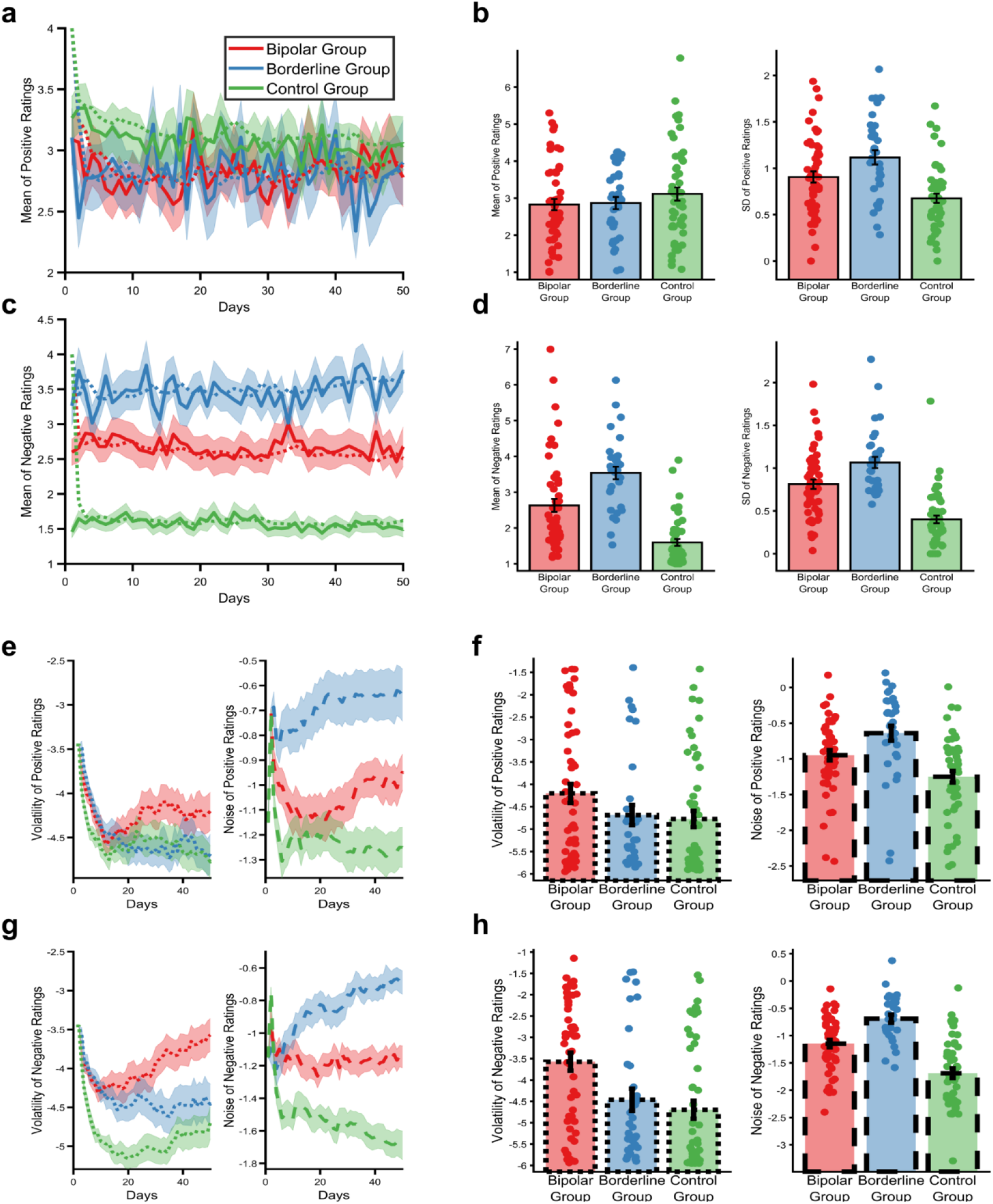
The types of affective variability in people with a diagnosis of bipolar disorder, borderline personality disorder or no diagnosis. Mean positive **(a)** and negative (**c**) daily affect ratings across 50 days of the study (solid lines illustrate means, ±SEM are represented by shaded regions). The predicted mean scores of the Bayesian filter (i.e. the expected values of *mu*_*t*_ before observing that day’s rating) are superimposed as dashed lines. Summary statistics (mean and standard deviation) of positive **(b)** and negative **(d)** affect ratings calculated across the 50 days. As can be seen, positive affect ratings did not differ between groups, whereas ratings of negative affect differed significantly with the borderline personality disorder group reporting the highest scores, followed by the bipolar disorder group and then the control group. The same ordering of groups was found for affective variability of both positive and negative affect as estimated by the standard deviation of the ratings (11). Evolution of the Bayesian filter’s beliefs about the causes of positive (**e)** and negative **(g)** affective variability across the same 50 days of the study. Lines represent the mean (±SEM) of the expected values of the *vmu*_*t*_ node, for volatility, and *SD*_*t*_ node, for noise. Final beliefs of the Bayesian filter, i.e. at day 50, about the types of variability for positive (**f**) and negative (**h**) affect. The filter attributes different types of affective variability to the two clinical groups, with noise being higher in the borderline personality disorder group and volatility in the bipolar group.

Applying the Bayesian filter to this data (Figure 2, panels e-h) provided clear evidence of a specific association between distinct types of affective variability and diagnosis [group x type of variability; *F*(2,122) = 7.92, *p* = 0.001], which did not differ between positive and negative ratings [group x cause of variability x valence; *F*(2,122) = 1.91, *p* = 0.15]. As can be seen, across both positive and negative ratings, estimated volatility was higher in the bipolar group than in both the borderline [*p*_*bonf*_ = 0.042] and control [*p*_*bonf*_ < 0.01] groups, with the difference between the borderline and control groups being non-significant [*p*_*bonf*_ = 0.6]. In contrast, estimated noise was higher in the borderline group than in both the bipolar [*p*_*bonf*_ = 0.016] and control [*p*_*bonf*_ < 0.001] groups and was also higher in the bipolar than control [*p*_*bonf*_ < 0.001] group. In other words, the filter-estimated types of affective variability are diagnostically specific, with volatility being higher in patients with bipolar disorder and noise higher in patients with borderline personality disorders.

### Ongoing Lithium Treatment is Associated with Increased Volatility of Positive Affect

Of the 51 patients with a diagnosis of bipolar disorder recruited to the cohort study, 22 were currently receiving ongoing lithium treatment and 29 were not. As illustrated in Figure 3a,c, the volatility of positive ratings was raised in patients with a diagnosis of bipolar disorder who were receiving lithium treatment compared to those who were not [*F*(1,41)=6.27, *p*=0.023], with no difference in any of the other filter derived metrics [all *F*’s < 0.019, *p*’s > 0.89] and no difference in the mean or standard deviation of the ratings [all *F*’s < 1.425, *p*’s > 0.51]. Including lithium treatment as a factor in the analysis of the volatility data from the cohort study indicated that levels of positive volatility did not differ between the groups [main effect of group *F*(2,121)=0.35, *p*=0.71], but was influenced by lithium treatment status [main effect of lithium *F*(1,121)=4.66, *p*=0.033]. In contrast, negative volatility differed between the groups [main effect of group *F*(2,121)=7.14, *p* = 0.001] and was not influenced by lithium [main effect of lithium *F*(1,121) = 0.04, *p* = 0.84]. These results raise the possibility that lithium treatment increases the volatility of positive affect, although the design of this longitudinal study does not permit firm conclusions as to the causal effect of lithium treatment.

**Figure 3.**
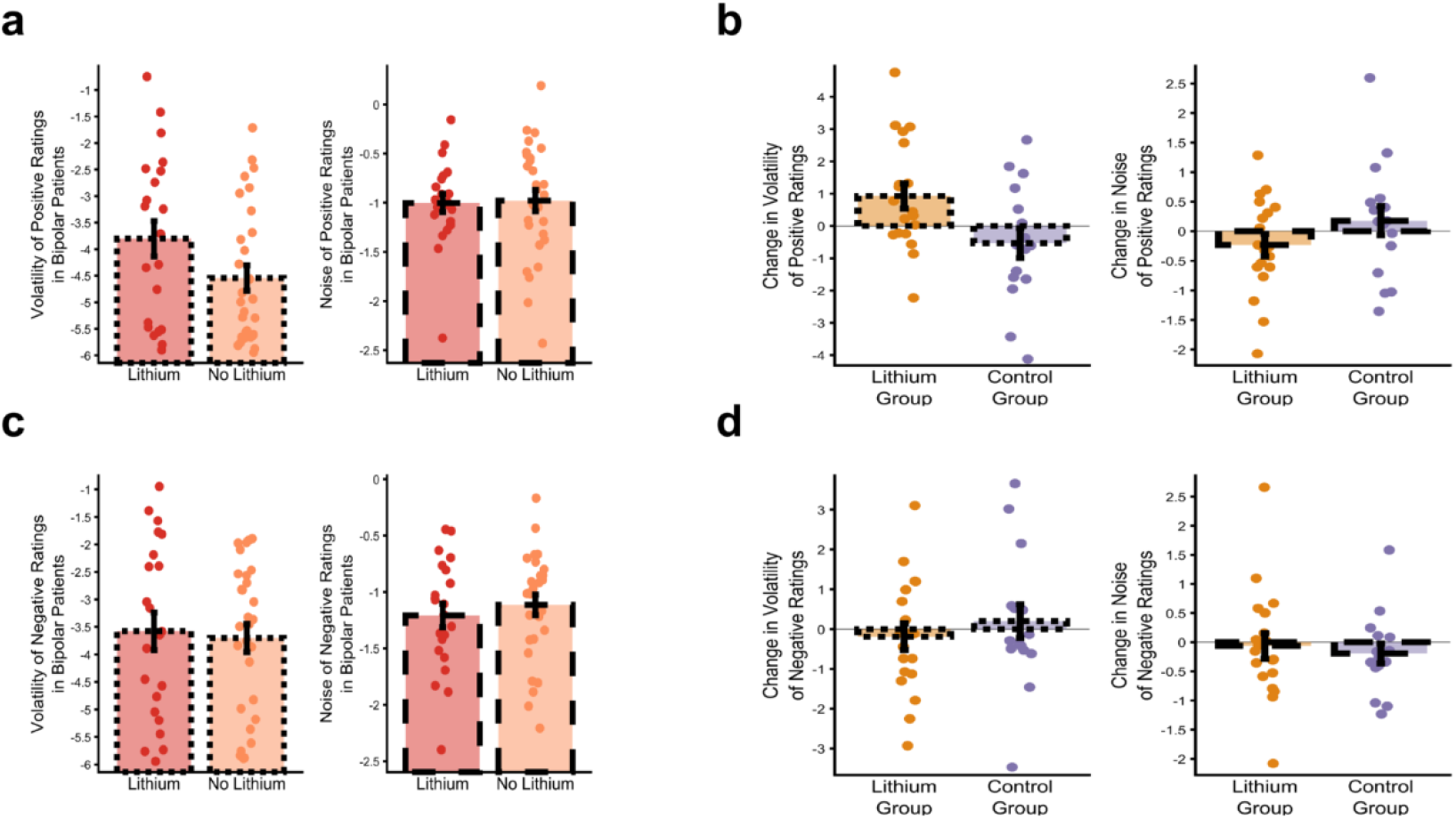
Lithium specifically increases the volatility of positive affect in patients with bipolar disorder. The volatility and noise of positive **(a)** and negative **(c)** affective ratings in patients from the bipolar group of the cohort study who were and were not receiving treatment with lithium. The charts illustrate the mean (±SEM) of volatility and noise at day 50. Patients receiving lithium have a higher volatility of positive affect (panel **a**), with no effect on the noise of positive ratings or on either outcome for negative ratings (panels **a, c**). While this result raises the possibility that lithium causes an increase of positive affective volatility, strong evidence for causality requires a randomized design. Panels **b** and **d** illustrate the results of a randomized trial of lithium, with the change in the volatility and noise of positive (**b**) and negative (**d**) affective ratings across the treatment period of the study shown. The charts illustrate the mean (±SEM) of the changes in volatility and noise at day 28 (relative to the end of the run-in period). As can be seen, the results of this randomized study are consistent with those from the cohort study, with lithium producing a specific increase in the volatility of positive affect ratings, with no effect on the other measures.

### Initiation of Lithium Specifically Increases Positive Affective Volatility in Bipolar Disorder

The causal influence of Lithium on affective variability was therefore assessed using data from the Oxford Lithium (OxLith) trial (22). In this study, patients with a diagnosis of Bipolar Disorder were randomly assigned to 4 weeks treatment with lithium or placebo, with daily affect ratings completed from 2 weeks before treatment initiation to 4 weeks after. Lithium treatment had no effect on the means [all interactions including treatment; *F*’s(1,33) < 1.6, *p*’s > 0.21] or standard deviations [*F*’s(1,33) < 0.67, *p*’s > 0.42] of either the positive or negative affect ratings. However, lithium differentially altered affective variability, estimated using the Bayesian filter, as a function of both its type and valence [treatment x type of variability x valence *F*(1,33)=5.68, *p* = 0.02]. As can be seen in Figure 3b,d and consistent with the results from the cohort study (Figure 3a,c) lithium specifically increased the volatility of positive affect ratings [effect of treatment *t*(33)=2.17, *p* = 0.04], without altering negative volatility [*t*(33)=-0.9, *p* = 0.39] or the noise of either valence [*t’*s(33) < 1.36, *p*’s > 0.18].

## Discussion

Patients with bipolar disorder and borderline personality disorder were found to have distinct types of affective variability, as defined by a generative model of affect. Affective volatility was increased in patients with bipolar disorder, whereas affective noise was increased in patients with borderline personality disorder. Treatment with lithium specifically elevated the volatility of positive affect.

There has been debate about the types of affective variability that might exist, how the different types should be defined and whether they add to simpler metrics such as the mean and standard deviation of affective ratings (11–14). The measures derived from our model indicate, for the first time, specificity of the type of variability of affect ratings between patients with diagnoses of bipolar and borderline personality disorders. This observed association, with increased affective volatility in patients with bipolar disorder and increased affective noise in patients with borderline personality disorder is consistent with the qualitative descriptions of the disorders (4) and indicates that the model formally captures clinically relevant aspects of affective variability that are not apparent using simpler metrics (2, 11, 15).

The Bayesian filter estimates different forms of variability within a single framework. A range of alternative measures of affective variability have previously been described (14). Of these, the measure most closely linked to affective volatility is affective inertia, often formalized as the slope of a first degree autoregressive (AR1) model (23, 24), with affective noise being similar to the standard deviation of the residuals of that model. Previous work has associated increased affective inertia with reduced functioning (23, 24) and analysis of the current data using an AR1 model produced a similar overall pattern of results for the cohort study, although did not replicate the difference between the bipolar and borderline clinical groups found for volatility (see supplementary information).

We found that lithium, an agent with proven efficacy for treating and averting extreme affective states in bipolar disorder (9, 25), specifically increased the volatility of positive affect of patients with bipolar disorder in both a real-world cohort study and a randomized experimental medicine study. A question raised by these results is how an increase in positive affective volatility might relate to the clinical effects of lithium, particularly its ability to terminate or avoid extreme mood states (25). One explanation relates to a characteristic feature of mania and depression; that, during an episode, patients’ affect becomes stuck at an extreme. Affective volatility is a change in affect that persists across time, suggesting that increased positive volatility may be exactly what is required to escape the affective confines of manic or depressed episodes. In other words, lithium does not act to simply suppress affective variability, as might be assumed of a “mood stabilizing” treatment, but rather to enhance a particular type of affective variability which can prevent patients becoming stuck in periods of mania or depression. This interpretation raises a number of questions for future study, most obviously whether the clinical impact of lithium is related to its effect on positive volatility and whether other interventions that target affective noise and negative volatility may be identified.

The Bayesian filter attributes changes in affect that persist across sampling points to volatility, whereas changes that do not persist are attributed to noise (see supplementary materials). This suggests an interpretation of the current results in terms of the “half-life” of affective responses: Patients with bipolar disorder have a longer half-life of affective response than patients with borderline personality disorder (at least for negative affect) and lithium acts to increase the half-life of positive affective responses. This formulation is consistent with previous qualitative descriptions of these patient groups that have highlighted the shorter time scale of affective responses in patients with a diagnosis of borderline personality disorder (26) and suggests that the filter derived metrics may provide a particularly useful quantitative assessment of patients who lie at the diagnostic boundary, such as those with rapidly-cycling bipolar disorder (26).

An important limitation of this work relates to the characteristics of the cohorts used in the first study. In this study, the control group was specifically matched to the bipolar, rather than the borderline group. As a result, patients in the borderline group had a lower educational achievement and were much less likely to be male. While the same pattern of results was found when analyzing only female patients (see supplementary results) and these demographic variables were included in the analyses, it would clearly be desirable to replicate the current findings in separate cohorts in which these demographic factors are more closely matched.

Computational psychiatry uses formal descriptions of mechanistic processes to better understand psychiatric illness and enhance the development of novel treatments (27). Taking this approach, we have deployed a generative model and associated Bayesian filter to describe and measure distinct types of affective variability in patients with bipolar and borderline personality disorders, and have found that lithium acts to specifically increase positive affective volatility.

## Materials and Methods

Details of the cohort and experimental medicine studies are provided in sequential sections.

### Cohort study

#### Overview

The Automated Monitoring of Symptom Severity (AMoSS) study recruited cohorts of patients diagnosed with bipolar disorder and borderline personality disorder as well as non-clinical participants, in order to examine the relationship between affect, activity and physiological measures. Results from the AMoSS study, including summary measures of affect ratings and validation of the ratings used have been previously reported by Tsanas and colleagues (11). All participants gave written informed consent to participate in the study which was approved by the East of England, Norfolk NHS ethics committee (13/EE/0288). Participants were asked to complete three months of daily ratings of positive and negative affect with the option to continue indefinitely beyond this point. Participants were also asked to provide demographic data including age, sex and educational attainment.

#### Participants

Patients were recruited from services in Oxfordshire and from the local community. Control participants were gender and age matched to patients from the bipolar group. All participants were assessed by a consultant psychiatrist who confirmed diagnoses of bipolar disorder using the structured clinical interview for DSM-IV (SCID-IV)(28) and of borderline personality disorder using the appropriate section of the International Personality Disorder Examination (IPDE)(29). Control participants were screened using the SCID to confirm no current or previous diagnosis.

A total of 53 patients diagnosed with bipolar disorder, 33 diagnosed with borderline personality disorder and 53 control participants were recruited to the study. 1 participant withdrew consent. For the current analysis participants were included if they had completed at least 10 affect ratings (as estimates of volatility stabilized at this point, see Figure 1). This left 51 patients with bipolar disorder, 33 patients with borderline personality disorder and 51 controls. As the control group were selected to match the bipolar group, the group of patients with borderline personality disorder differed from the other two groups with a higher proportion of female patients and lower average educational attainment These variables were included as covariates in the reported statistics (and analysis restricted only to female participants produced the same group x type of affective variability effect; *F*(2,84)=3.1, *p*=0.05).

#### Measure of affect

Participants completed daily affect ratings using the “moodzoom” android app (participants without an android phone were supplied with one for the duration of the study). The moodzoom app prompted participants to rate their current affect every evening by endorsing each of six descriptors (anxious, elated, sad, angry, irritable, energetic) on a 7 point Likert scale. Summary positive and negative affect scores were calculated as the average of the positive and negative items(11). The 50 day period for each participant that had the fewest missing data points was used for analysis (identical group effects were observed if the first 50 ratings for each participant were used).

### Experimental medicine study

#### Overview

The Oxford Lithium (OxLith) trial was a randomized, controlled experimental medicine study, of patients with bipolar spectrum disorder, conducted in Oxford (22). During an initial screening visit diagnosis was confirmed using the SCID-IV, participants then completed a two-week, pre-randomization, run-in period, following which they were randomized to receive lithium carbonate or placebo for up to six weeks. Participants completed daily ratings of positive and negative affect throughout the run-in and post randomization periods. All participants provided written informed consent to participate in the study which had been approved by the NHS South Central Research Ethics Committee (15/SC/0109). The study protocol was registered (ISRCTN91624955) and published(22) before study completion. The analysis reported in the current paper is an additional exploratory analysis not described in the protocol.

#### Participants

The study recruited individuals aged 18 years or over, with a diagnosis of bipolar disorder (bipolar I, II or NoS) for whom there was uncertainty about whether treatment with lithium was appropriate (e.g. an individual with a recent diagnosis of bipolar disorder or who has experienced relatively few severe mood episodes). Individuals were recruited from local clinical services. Individuals were not eligible for the trial if they had any contraindications to lithium treatment, were taking concomitant psychotropic medication that they were unable to discontinue, had clinically significant substance misuse, required urgent treatment for a mood disorder (i.e. where placebo treatment would be unethical), were pregnant or of child bearing age and not using effective contraception or were acutely suicidal. Summary demographic data are presented in Table 1.

#### Randomization, intervention and blinding

Participants were randomized using a 1:1 allocation scheme which was minimized for participant age (<25 years, ≥25 years) and sex (female, male). The active group received lithium carbonate 200mg prolonged release tablets which was titrated to a target serum level of 0.7mmol/L as per routine practice. The trial psychiatrist and participants remained blind to treatment allocation. For participants in the placebo group, sham lithium levels were provided to the treating psychiatrist who then adjusted the placebo “dose”.

#### Measure of affect

Participants completed an online daily version of the positive and negative affect scale, 10 item version, (PANAS)(30). The PANAS requires participants to rate five positive descriptors (alert, inspired, determined, attentive, active) and five negative descriptors (upset, hostile, ashamed, nervous, afraid) on a five-point scale. Summary positive and negative affect ratings were calculated as the average of the positive and negative ratings respectively.

### The Bayesian Filter

Here we provide a summary of the generative model of affect and associated Bayesian Filter. A formal description, and a comparison with alternative models/measures, is provided in the supplementary materials. In the generative model (Figure 1a), one rating per time point (*y*_*t*_) is drawn from a Gaussian probability distribution with a mean, *mu*_*t*_, and a standard deviation, *SD*_*t*_, (Figure 1b). The mean can change between time points, with this change controlled by the volatility parameter, *vmu*_*t*_. Two higher level parameters, *kmu* and *vSD*, control the change over time of the volatility and standard deviation respectively, allowing the model to account for periods during which the volatility and/or standard deviation are high and periods when they are low. The generative model defines two causes of variability of the ratings (Figure 1b)(19): First, a change in the mean of the distribution between trials can cause variability in the ratings (e.g. if the mean has decreased then the ratings of the next trials will, on average, be lower). The size of this variability is controlled by the volatility parameter, *vmu*_*t*_. Second, the production of the ratings from a Gaussian distribution leads to variability about the mean that influences the current rating but has no carry over effects. The size of this variability, which we call noise, is controlled by the standard deviation, *SD*_*t*_, of the distribution.

The Bayesian filter inverts this generative model. It starts with the affect ratings, *y*_*t*_ and uses these to recursively update its belief about the state of the five generative processes (the circles above *y*_*t*_ in Figure 1a) which cause the ratings. As a result, the filter estimates, for each point in time, the degree to which the variability in ratings is produced by volatility and the degree to which it is produced by noise (Figure 1d).

Where more than one set of ratings were provided in a day the first was used, days in which no ratings were provided were treated as missing with no data extrapolation (see supplementary materials for an illustration of how the Bayesian filter deals with missing data and for sensitivity analysis of data missingness).

### Statistical Analysis

Analysis of filter-based data from the cohort study was performed using repeated measures ANOVAs with the within subject factors of cause of variability (volatility, noise) and valence (positive, negative) and the between subject factor of group (bipolar group, borderline group, control group). In addition, age, gender and educational attainment were included as control variables in all analyses. In these analyses the dependent variables were the filter derived estimates of volatility and noise at day 50. Post hoc comparison between the groups was carried out with Bonferroni correction and is labelled as such in the paper. The filter-based data was not normally distributed and so was boxcox transformed (lambda=0.2) before entry into the analysis. As demographic data were missing from some participants (see Table 1) the reported statistical analyses are limited to participants with complete demographic data (omission of these control variables and inclusion of all participants in the analysis or analysis of the untransformed data does not alter the significance of results). Data from all participants are included in the figures. The nonfilter-based metrics (mean and standard deviation) were analyzed separately (as the mean is not a cause of variability) with a single within subject variable of valence.

Analysis of data from the experimental medicine study was carried out using a repeated measures ANOVA with the within subject factors of cause of variability (volatility, noise), and valence (positive, negative) and the between subject factors of group (lithium, placebo). The dependent variables used were the change in filter derived estimates of variability between the end of the run-in period and the end of the treatment period. These data did not violate normality assumptions and so were not transformed. All inferential statistical tests were two-sided. Analyses were performed using IBM SPSS version 25.

## Supporting information

Supplementary Methods and Results

## Data Availability

The cohort and experimental medicine studies did not obtain consent to upload data onto open platforms.
However anonymous data can be shared with other research groups who have ethical approval in place on a by-project basis. Requests for data access should be made to

## Acknowledgments

MB was supported by a MRC Clinician Scientist Fellowship (MR/N008103/1). MB, JG, CJH, and KEAS are supported by the Oxford Health NIHR Biomedical Research Centre. The views expressed are those of the authors and not necessarily those of the NHS, the NIHR or the Department of Health. The cohort study was supported by the Wellcome Trust through a Centre Grant (no. 98,461/Z/12/Z, The University of Oxford Sleep and Circadian Neuroscience Institute (SCNi)) and by a Wellcome Trust Strategic Award (CONBRIO: Collaborative Oxford Network for Bipolar Research to Improve Outcomes, Reference number 102,616/Z) which also funded the experimental medicine study.

## References

1. H. W. Koenigsberg, Affective instability: toward an integration of neuroscience and psychological perspectives. J. Pers. Disord. 24, 60–82 (2010).

2. C. Henry, et al., Affective instability and impulsivity in borderline personality and bipolar II disorders: similarities and differences. J Psychiatr Res 35, 307–312 (2001).

3. C. Henry, et al., Affective lability and affect intensity as core dimensions of bipolar disorders during euthymic period. Psychiatry Research 159, 1–6 (2008).

4. D. B. Reich, “Affective Instability: Bipolar Disorder Versus Borderline Personality Disorder” in Borderline Personality and Mood Disorders: Comorbidity and Controversy, L. W. Choi-Kain, J. G. Gunderson, Eds. (Springer, 2015), pp. 79–95.

5. S. Marwaha, N. Parsons, M. Broome, Mood instability, mental illness and suicidal ideas: results from a household survey. Soc Psychiatry Psychiatr Epidemiol 48, 1431–1437 (2013).

6. S. Marwaha, M. R. Broome, P. E. Bebbington, E. Kuipers, D. Freeman, Mood instability and psychosis: analyses of British national survey data. Schizophr Bull 40, 269–277 (2014).

7. S. Marwaha, et al., How is affective instability defined and measured? A systematic review. Psychological Medicine 44, 1793–1808 (2014).

8. M. R. Broome, K. E. A. Saunders, P. J. Harrison, S. Marwaha, Mood instability: significance, definition and measurement. Br J Psychiatry 207, 283–285 (2015).

9. J. R. Geddes, S. Burgess, K. Hawton, K. Jamison, G. M. Goodwin, Long-term lithium therapy for bipolar disorder: systematic review and meta-analysis of randomized controlled trials. Am J Psychiatry 161, 217–222 (2004).

10. J. Stoffers, et al., Pharmacological interventions for borderline personality disorder. Cochrane Database of Systematic Reviews (2010) https://doi.org/10.1002/14651858.CD005653.pub2 (May 20, 2020).

11. A. Tsanas, et al., Daily longitudinal self-monitoring of mood variability in bipolar disorder and borderline personality disorder. J Affect Disord 205, 225–233 (2016).

12. E. L. Hamaker, E. Ceulemans, R. P. P. P. Grasman, F. Tuerlinckx, Modeling Affect Dynamics: State of the Art and Future Challenges. Emotion Review 7, 316–322 (2015).

13. T. J. Trull, S. P. Lane, P. Koval, U. W. Ebner-Priemer, Affective Dynamics in Psychopathology. Emotion Review 7, 355–361 (2015).

14. E. Dejonckheere, et al., Complex affect dynamics add limited information to the prediction of psychological well-being. Nat Hum Behav 3, 478–491 (2019).

15. H. W. Koenigsberg, et al., Characterizing affective instability in borderline personality disorder. Am J Psychiatry 159, 784–788 (2002).

16. P. Santangelo, et al., Specificity of Affective Instability in Patients With Borderline Personality Disorder Compared to Posttraumatic Stress Disorder, Bulimia Nervosa, and Healthy Controls. J Abnorm Psychol 123, 258–272 (2014).

17. E. A. Selby, et al., Momentary emotion surrounding bulimic behaviors in women with bulimia nervosa and borderline personality disorder. J Psychiatr Res 46, 1492–1500 (2012).

18. T. E. J. Behrens, M. W. Woolrich, M. E. Walton, M. F. S. Rushworth, Learning the value of information in an uncertain world. Nat. Neurosci. 10, 1214–1221 (2007).

19. E. Pulcu, M. Browning, The Misestimation of Uncertainty in Affective Disorders. Trends Cogn. Sci. (Regul. Ed.) (2019) https://doi.org/10.1016/j.tics.2019.07.007.

20. R. E. Kalman, A New Approach to Linear Filtering and Prediction Problem. Transactions of the ASME 82, 35–45 (1960).

21. C. D. Mathys, et al., Uncertainty in perception and the Hierarchical Gaussian Filter. Front Hum Neurosci 8, 825 (2014).

22. K. E. A. Saunders, et al., Oxford Lithium Trial (OxLith) of the early affective, cognitive, neural and biochemical effects of lithium carbonate in bipolar disorder: study protocol for a randomised controlled trial. Trials 17, 116 (2016).

23. P. Koval, S. Sütterlin, P. Kuppens, Emotional Inertia is Associated with Lower Well-Being when Controlling for Differences in Emotional Context. Frontiers in Psychology 6 (2016).

24. P. Kuppens, N. B. Allen, L. B. Sheeber, Emotional Inertia and Psychological Maladjustment. Psychol Sci 21, 984–991 (2010).

25. A. Cipriani, et al., Comparative efficacy and acceptability of antimanic drugs in acute mania: a multiple-treatments meta-analysis. Lancet 378, 1306–1315 (2011).

26. D. F. MacKinnon, R. Pies, Affective instability as rapid cycling: theoretical and clinical implications for borderline personality and bipolar spectrum disorders. Bipolar Disorders 8, 1–14 (2006).

27. M. Browning, et al., Realizing the Clinical Potential of Computational Psychiatry: Report From the Banbury Center Meeting, February 2019. Biol. Psychiatry 88, e5–e10 (2020).

28. M. B. First, R. L. Spitzer, M. Gibbon, J. B. W. Williams, Structured Clinical Interview for DSM-IVTR Axis I Disorders, Research Version, Patient Edition. (SCID-I/P) (New York State Psychiatric Institute, 2002).

29. A. W. Loranger, et al., The International Personality Disorder Examination: The World Health Organization/Alcohol, Drug Abuse, and Mental Health Administration International Pilot Study of Personality Disorders. Arch Gen Psychiatry 51, 215–224 (1994).

30. D. Watson, L. A. Clark, A. Tellegen, Development and validation of brief measures of positive and negative affect: the PANAS scales. J Pers Soc Psychol. 54, 1063–1070 (1988).

