## Supplementary Methods and Results for "Characterizing Affective Variability in Bipolar Disorder and Borderline Personality Disorder, and the Effects of Lithium, Using a Generative Model of Affect"

In the supplementary materials we provide a formal description of the Bayesian filer, explore the effect of alternative link functions, assess the performance of the filter and describe a series of sensitivity and exploratory analyses that assess potential confounding factors and alternative explanations for the results reported in the main paper.

**Supplementary Methods**

The Bayesian Filter

*Overview:* A grid-based recursive Bayesian filter(1) was developed to estimate the causal processes described in the generative model (Figure 1). Note that the filter has no free parameters and so is not fitted to participant ratings, rather it uses the ratings to estimate the values of the causal processes defined by the generative model.

As illustrated in Figure 1a, the filter assumes that affect ratings at a given time point $t$, $y_{t}$, are generated from a Gaussian distribution with an unknown mean, ${mu}_{t}$, and standard deviation, $exp({SD}_{t})$, with the later producing noise in the ratings (Figure 1b). Note that all variance parameters in the filter are represented in log space to ensure they lie on the infinite real line. Formally, the production of ratings is described by:

$y_{t}\sim\mathcal{N(}{mu}_{t},\exp\left( {SD}_{t} \right))$	1

The mean of this distribution may change between time points, leading to volatility of the ratings (Figure 1b), with this change described by a second Gaussian distribution, centered on the current mean and with a standard deviation of $\exp({vmu}_{t})$.

${p(mu}_{t+1})\sim\mathcal{N(}{mu}_{t},\exp\left( {vmu}_{t} \right))$	2

Both the ${SD}_{t}$ and ${vmu}_{t}$ parameters can also change between time points with their change governed by Gaussian distributions centred on their current value with standard deviations of $\exp(vSD)$ and $\exp(kmu)$ respectively. These higher-level parameters allow the model to conceptualise periods in which noise and volatility are high and other periods in which they are low.

${p(vmu}_{t+1})\sim\mathcal{N(}v{mu}_{t},\exp\left( kmu \right))$	3

${p(SD}_{t+1})\sim\mathcal{N(}{SD}_{t},\exp\left( vSD \right))$	4

The filter seeks to estimate the joint posterior probability of the five causal parameters, given the affect ratings it has observed. The joint probability distribution at time point $t$ is defined as:

$p\left( {joint}_{t} \right)=p\left( {mu}_{t},{vmu}_{t},kmu,{SD}_{t}, vSD \right|y_{t-1}, y_{t-2},\ldots y_{1})$	5

This joint probability distribution can be thought of as the filter's belief about the values of each parameter in the generative model. The filter uses this distribution as a sufficient statistic of the affect generating process (i.e. it assumes that the magnitude of future affect ratings is completely described by this distribution). Formally, this is equivalent to a Markovian assumption (i.e. that the joint probability on the next trial depends only on the joint probability of this trial and the observed rating) so that that recursive update performed by the filter can be stated as:

$p\left( {joint}_{t} \right)=p\left( {mu}_{t},{vmu}_{t},kmu,{SD}_{t}, vSD \right|y_{t-1},{joint}_{t-1})$	6

We initialize the joint posterior, before observation of any ratings, $p({joint}_{1})$, as a flat distribution. In the next section we describe how the recursive updates are performed between time points.

*Belief update1: the effect of the observed rating*: The update of the joint probability distribution between time points is split into two broad components. First the filter has to use the rating it observes to update its belief. This is achieved using Bayes rule:

$p\left( {joint}_{t-1} \right| y_{t-1})=\frac{p\left( y_{t-1} \right|{joint}_{t-1}) p({joint}_{t-1})}{p(y_{t-1})}$	7

The important points to note here are 1) the likelihood term, $p\left( y_{t-1} \right| {joint}_{t-1})$, is a function only of the mean, ${mu}_{t}$, and standard deviation, ${SD}_{t}$, of the generative model and 2) the process described by this equation does not account for the change in values of the ${mu}_{t}$, ${vmu}_{t}$ or ${SD}_{t}$ parameters between time points described by equations 2-4.

*Belief update2: the effect of parameter shifts*: The next component of the update deals with the change in the ${mu}_{t}$, ${vmu}_{t}$ or ${SD}_{t}$ parameters across time. In order to illustrate this in an intuitive fashion, we first describe the update of a single parameter, ${mu}_{t}$, from time $t-1$ to time $t$, given the volatility, ${vmu}_{t}$.

$p\left( {mu}_{t} \right| {vmu}_{t})=\int p\left( {mu}_{t} \right| {mu}_{t-1}, {vmu}_{t}) d({mu}_{t-1})$	8

The integral in this equation sums together the probabilities, across every possible starting value of the mean,${mu}_{t-1}$, that would have led it now to be the new value, ${mu}_{t}$, as defined by $p\left( {mu}_{t} \right| {mu}_{t-1}, {vmu}_{t})$. In this way the original value of the mean is "integrated out" and the probability distribution across values of ${mu}_{t-1}$ is updated. The same process is applied to the other drifting parameters in the model and is combined with the update in response to the observed rating described in equation 7 to give:

$$p\left( {joint}_{t} \right|y_{t-1})=$$

$$\iiint p\left( {joint}_{t-1} | y_{t-1} \right)p\left( {SD}_{t} \right| {SD}_{t-1},vSD)p\left( {vmu}_{t} | {vmu}_{t-1},kmu \right)\ldots$$

$p\left( {mu}_{t} | {mu}_{t-1},{vmu}_{t} \right), d{SD}_{t-1}, d{vmu}_{t-1}, d{mu}_{t-1}$	9

The recursive nature of the filter and the separation of the updating process into two components provides a natural approach for dealing with missing data. On days when no rating was returned the first phase of the update (equation 7) is omitted, but the second phase (remaining terms in equation 9) is applied, leading to a dispersion of the filter's belief about the current parameter values (see supplementary methods for an illustration of this and for a sensitivity analysis).

The filter's belief about the value of each node is derived at every time point by marginalising over all but the relevant dimension of the joint probability distribution and calculating the expected value of that dimension.

Note that the filter estimates magnitudes in an unbounded space, whereas affect ratings are produced on a bounded scale (e.g. from 1-7). The magnitude ratings were therefore logit transformed before being passed to the filter. An alternative approach to this issue is to specify an alternative link function for the filter. In the next section we describe such a link function and show that it produces equivalent results.

The filter was implemented as a five dimensional matrix in matlab (R2020). The filter's code is available at: <https://osf.io/j7md3/>

**Supplementary Results**

An Alternative Link Function

While the filter described above acts on data points from the infinite real line, the affective ratings collected from participants are constrained between the upper and lower bounds of the scale used (e.g. the individual items from the MoodZoom Scale require discrete responses between 1-7, which are averaged into positive or negative scores). A logit transform of the data is therefore used before it is presented to the filter. An alternative approach to this issue is to specify a link function which explicitly describes how the ratings are generated. For ordered, discrete responses, one such link is the ordered-probit function (2). In brief, this assumes that a gaussian latent process underlies the (discrete, ordinal) data, but that the data are generated using a set of cut points of this function (e.g. if data take the form 1:n, then n-1 cut points are defined with samples below cut point one producing a data value of “1”, between cut points 1-2 producing a value of “2” etc). We incorporated a version of the ordered-probit link function to the Bayesian filter (details of function provided below). In figure S1, we show that the results using this link function on untransformed data (Figure S1 c-d) is similar to the filter described in the paper on the transformed data (Figure S1a-b). As can be seen, using the ordinal probit link function resulted in the same pattern of diagnostic specificity for affective variability (Figure S1c-d; Group x Type of Variability *F*(2,122)=7.84, *p*<0.001) as that reported in the main paper (data reproduced in Figure S1a-b), with the estimates of the two versions of the filter correlating at *r*>0.8 for volatility and at *r*>0.75 for noise. This suggests that the results reported in the main paper are not sensitive to the specific approach used to transform the data.

*Details of Ordinal Probit Link Function:* Ordinal probit functions have a number of different parameters that may be varied (e.g. the position of the cut points, the position and variance of the latent process). If all of these are allowed to vary simultaneously, the function is not identifiable (e.g. increasing the SD of the latent process is equivalent to moving the cut points closer together). This requires some of the parameters to be set to specific values. For example, for analyses of stationery processes the value of the lower cut point is often set to 0, and the SD of the latent process is set to 1. In the current analyses we are interested in how the SD and the variability of the mean of the latent process differ between groups and therefore cannot fix these. We therefore fix the positions of the cut points to the set values of 1,2,3,...n-1. This is implemented in the filter by adjusting the calculation of the likelihood (Equation 7), with the likelihood of a data point between two cut points (e.g. lying in the range 1-2) being the total probability density of the latent process between these two values.

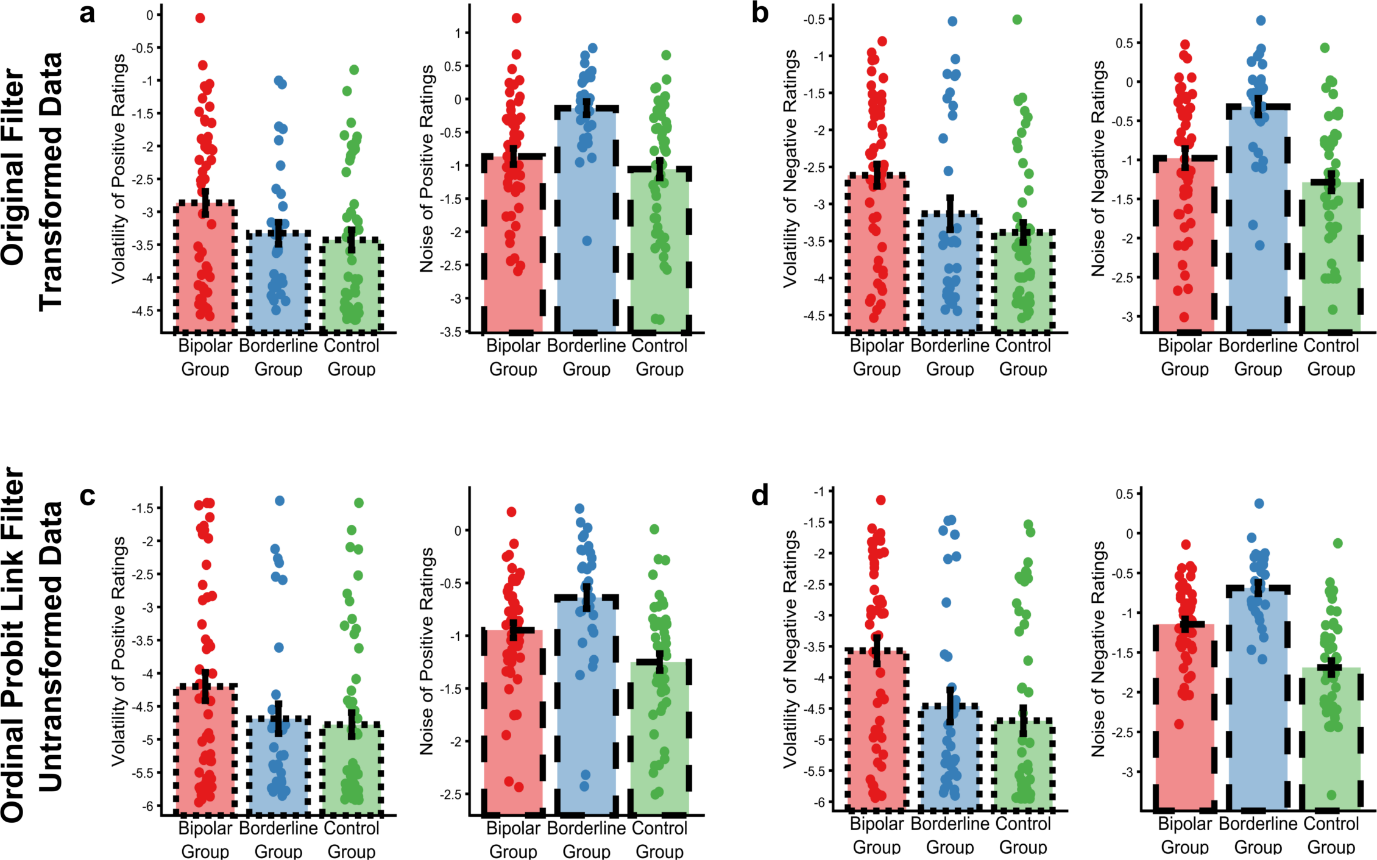

Figure S1: Estimated types of affective variability from the cohort study when the Bayesian filter is applied to transformed ratings data (panels a-b, reproduced from the main paper) and when an ordinal probit link function is used, so that the filter may be applied to untransformed data (panels c-d). As previously, the volatility and noise for the positive and negative ratings are presented separately. Both versions of the filter estimate volatility to be raised in patients with a diagnosis of bipolar disorder, with noise raised in patients with a diagnosis of borderline personality disorder.

Performance of the Filter

The ability of the filter to recover parameters, when those parameters are drifting across time, was assessed using a generate recover procedure. The generative model described in main Figure 1 was used to produce 90 sets of artificial data. This was achieved by selecting a starting state of the generative model by randomly sampling the state of each model node from a Gaussian distribution with mean and SD set to those of the cohort sample of participants. The generative model was then allowed to run for fifty time points (“days”), with the values of the mean, volatility and noise drifting between time points as described by the generative model (note that the other nodes do not drift). Synthetic affective ratings were sampled at each time point from the model’s mean and SD on that day. These data were then fed back to the Bayesian filter. The performance of the filter was assessed at day fifty (i.e. the same timepoint used in the study), by correlating the actual value of each drifting parameter with that recovered by the filter. The results of this process are illustrated in Figure S2. As can be seen, all three parameters are generally well recovered, with very low levels of volatility being the least accurately reproduced. This suggests that the filter is able to estimate these dynamic processes with a reasonable precision.

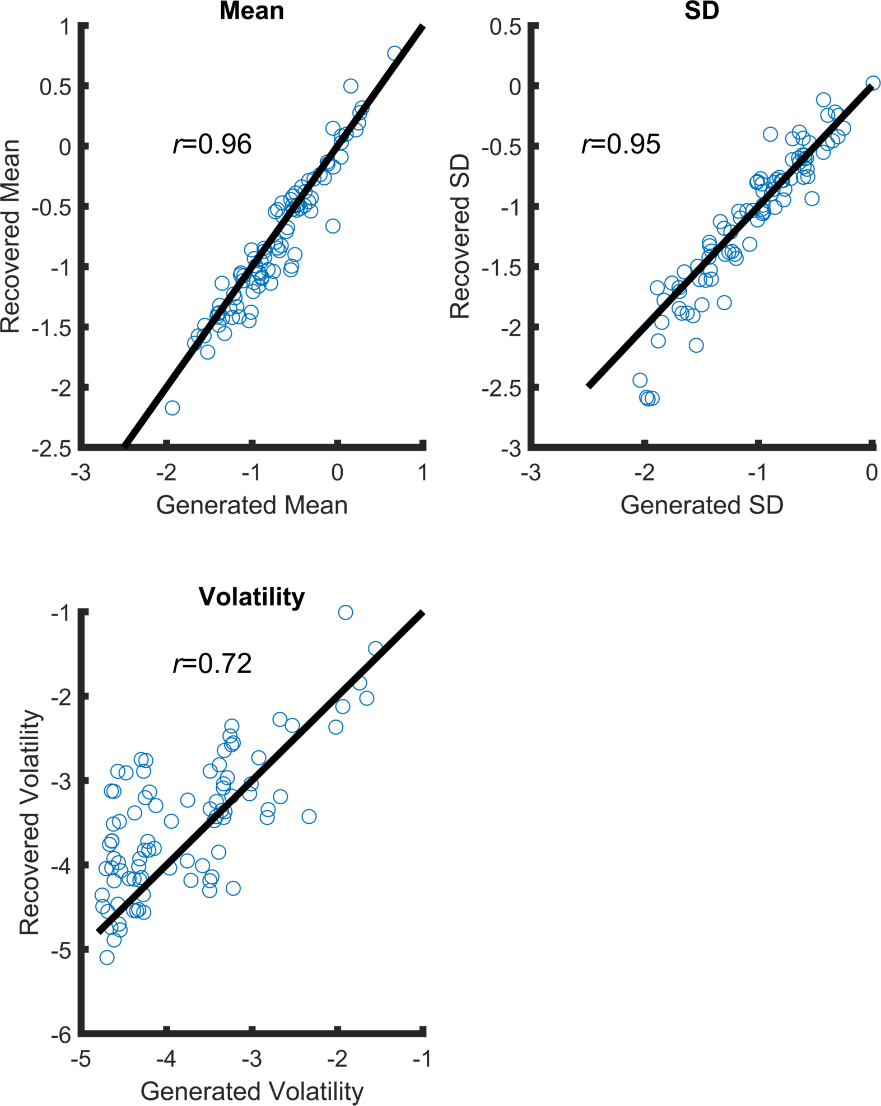

Figure S2: Results of a generate recover process. 50 days of artificial data were created for 90 pseudo-participants using the generative model described in Figure 1. The starting state of the model was set using the values from study participants, with the values of the mean, noise and volatility nodes allowed to drift over time as specified by the generative model. The filter was applied to the data generated using this process, with the filter-based estimates of mean, noise and volatility on day 50 correlated with the actual values of these parameters. As can be seen, the recovered values for the three parameters track the true underlying values. Black lines on each plot are y=x, that is, the line of perfect recovery.

Comparison of the filter with other measures of affective variability

A range of other measures of affective variability have been described (3). The measures which are most closely linked to those described in the current paper are affective inertia, commonly defined as the slope of an AR1 model, which is similar to the volatility term of the filter, and the standard deviation of the residual data, having fit the AR1 model, which is similar to the noise of the filter. Deriving the individual slopes from an AR1 model applied to data from the cohort study, as well as the standard deviation of the residuals, confirmed the overall results from Bayesian Filter with a significant group x variability type interaction (*F*(2,114)=13.3, *p*<0.01; note that missing data, or low levels of variability on one of the scales prevented fitting of the AR1 model in 8 participants, and so reduced the size of the groups by 2 in the group of patients with bipolar disorder, 1 in the group of patients with borderline personality disorder and 5 in the control participants). Post hoc tests found a significant difference in AR1 slope between the bipolar and control groups (*p_bonf_*<0.009), although the difference between the bipolar and borderline groups found for volatility was not replicated (whether corrected or not for multiple comparisons). Similar to the volatility results, the AR1 slope did not differ between borderline and control groups. Group differences for the standard deviation of the residuals were consistent with those of noise from the filter, with all group differences being significant at *p_bonf_*<0.045.

Analysis of metrics from the AR1 model for the experimental study was limited as missing rating data prevented the AR1 model being applied in five participants. The overall group x type of variability x valence effect here was not significant *F*(1,28)=2.17, *p*=0.15.

Overall these results indicate a degree of similarity between the filter derived measures and those obtained using an AR1 model, although there are differences, particularly in how the two approaches deal with missing data (see below for further analysis of missingness).

Missing data

*Treatment of Missing Data by the Filter*: When the filter encounters missing data it omits the Bayesian update defined in equation 7 but does apply the shift in parameter values defined by the generative model, as described by the remaining terms in equation 9. The effect of missing data is therefore to cause the filter to become progressively less certain about the values of the generative parameters. The rate at which this reduction of certainty occurs is rationally influenced by the model's belief about how changeable the parameters are. This process is illustrated with synthetic data in Figure S3 panels a and b for the ${mu}_{t}$ parameter. The solid black lines in these plots are the data fed to the filter, which is either volatile (panel a) or stable (panel b). The data from time point 50 onwards is missing. Superimposed on the line is the marginal distribution of the filter's belief about ${mu}_{t}$. As can be seen, this belief is concentrated around the observed data. The key aspect of these figures is what happens to the filter's belief when it encounters missing data. In panel a, where the filter has learned that the data is volatile, its belief quickly disperses so it becomes rapidly uncertain about the likely value of ${mu}_{t}$. In contrast, when the filter has learned that the data is stable, it maintains its belief about the value of ${mu}_{t}$ for a longer period. Thus the filter appropriately adapts its belief about the generative parameters of the data it observes as a function of its estimates about the changeability of these parameters.

*Missing Data Sensitivity Analysis*: In the cohort study there was no overall group difference in the amount of missing data [*F*(2,107)=0.86, *p*=0.42]. We conducted an additional sensitivity analysis to investigate whether the group differences in types of affective variability might be caused by some difference in the distribution of data missingness. This analysis simply censored a participant's data as soon as that participant misses a day's rating. The results of this analysis are illustrated in Figure S3 panels c and d. As can be seen, while the removal of participants increases the estimated error of the mean (the number of participants remaining in the three groups by day 50 are: Bipolar group, 10; Borderline group, 13; Control group, 19), the same overall pattern of group differences remains apparent. In other words, the group differences in cause of affective variability cannot be attributed to differences in data missingness.

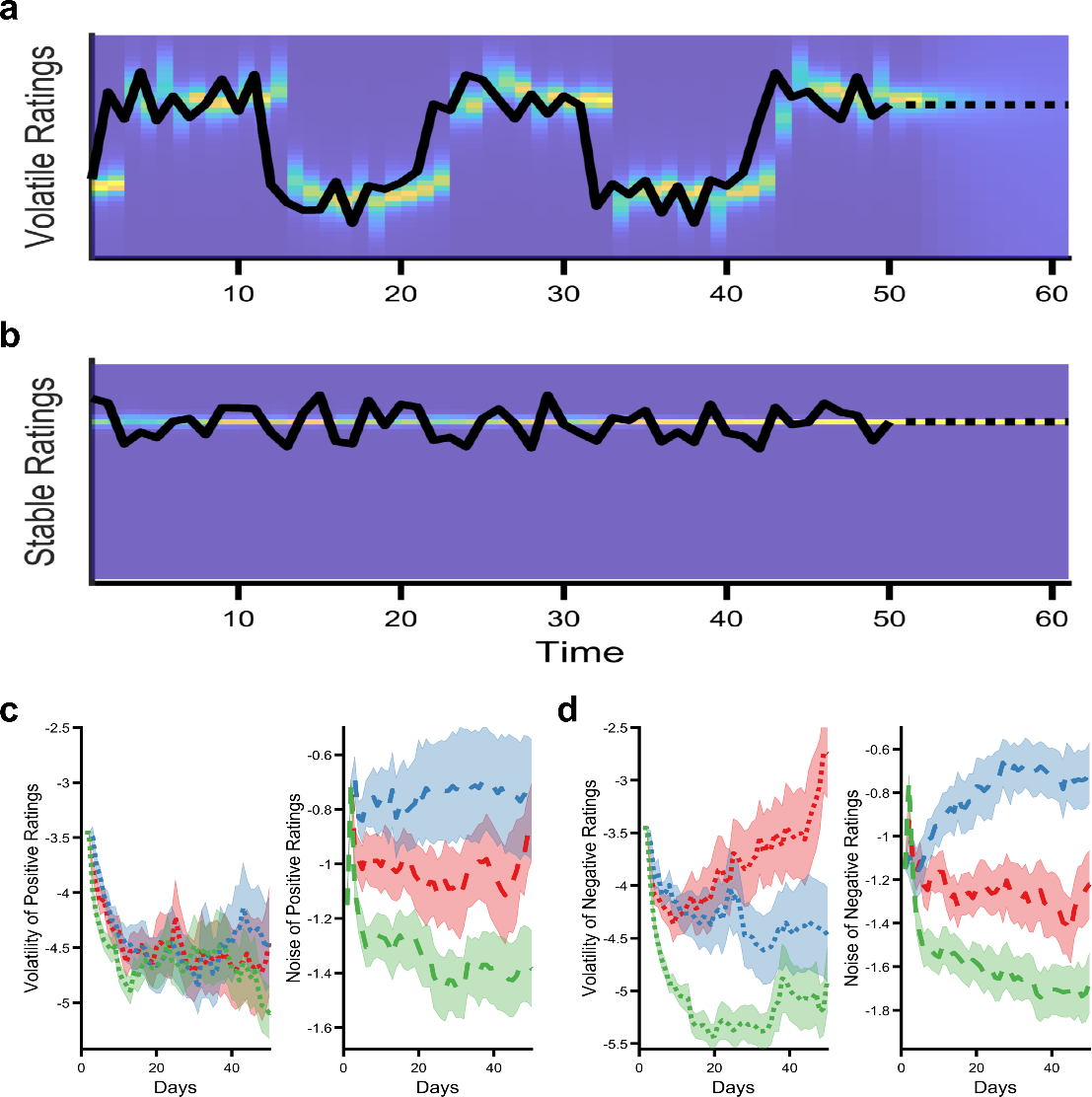

Figure S3: Effect of missing data on the estimated type of affective variability. The filter adapts rationally to missing data, dispersing its belief more quickly about parameters it believes to be changeable e.g. the ${mu}_{t}$ parameter when it has been volatile as shown in panel **a** compared to when it is stable as shown in panel **b**. See text for detailed explanation. Solid black lines indicate synthetic data fed to the filter, dotted lines are missing data. The background colour illustrates the filter's marginal belief about ${mu}_{t}$ which disperses more quickly in the volatile than stable example. Data missingness does not account for group differences in the cohort study. Panels **c** and **d** show the evolution of the filter's belief about the causes of affective variability as previously shown in Figure 2. In the current figure a participant's data is censored as soon as a single day's data is missing. The same pattern of group differences are apparent indicating that this effect is not related to differences in data missingness.

Duration of affective responses

The filter attributes changes in affect that persist across ratings to volatility and changes that do not to noise. This process is illustrated, using synthetic data, in Figure S4 panels a and b. Synthetic timeseries of affect ratings were generated by applying a series of perturbations that caused the affect rating to change, with this change gradually wearing off over time. These perturbations were intended to mimic the effect of events occurring in a patient's life that induced a change in affect. We varied both the frequency with which the perturbations occurred and the rate at which their effect wore off. Characteristic time courses of events at the four extremes (i.e. rare vs. frequent events, brief effect on ratings vs. sustained effect) are illustrated at the appropriate corners of Figure S4 panels a and b. The synthetic data was then passed to the filter which estimated the volatility (panel a) and noise (panel b) of the timeseries. As can be seen, estimates of both volatility and noise increase as the frequency of perturbations increase, whereas the duration of the perturbation influences the attribution of the variability, with long lasting perturbations being attributed to volatility and short lasting perturbations being attributed to noise.

This suggests that patients with bipolar disorder may have a longer duration of affective response than patients with borderline personality disorder (at least for negative affect) and that lithium acts to increase the duration of positive affective responses. In the main study we analysed daily affect ratings and therefore cannot assess timescales shorter than this. However, a significant proportion of participants in the cohort study provided daily ratings for longer than a year, allowing us to assess whether the observed group differences in affective variability are limited to data sampled once a day or whether they are also apparent at longer timescales. The results of these analyses are illustrated in Figure S4 panels c-f. In these analyses the first rating returned every week (panels c-d) or every three weeks (panels e-f) were passed to the filter. As can be seen the group differences in cause of affective variability remains apparent at the longer time periods. These results suggest that the duration of shifts of negative affective in the bipolar group have a duration of at least three weeks. An outstanding question concerns the duration of shifts in the borderline group, assessing this would require collecting affect ratings at a frequency greater than once daily.

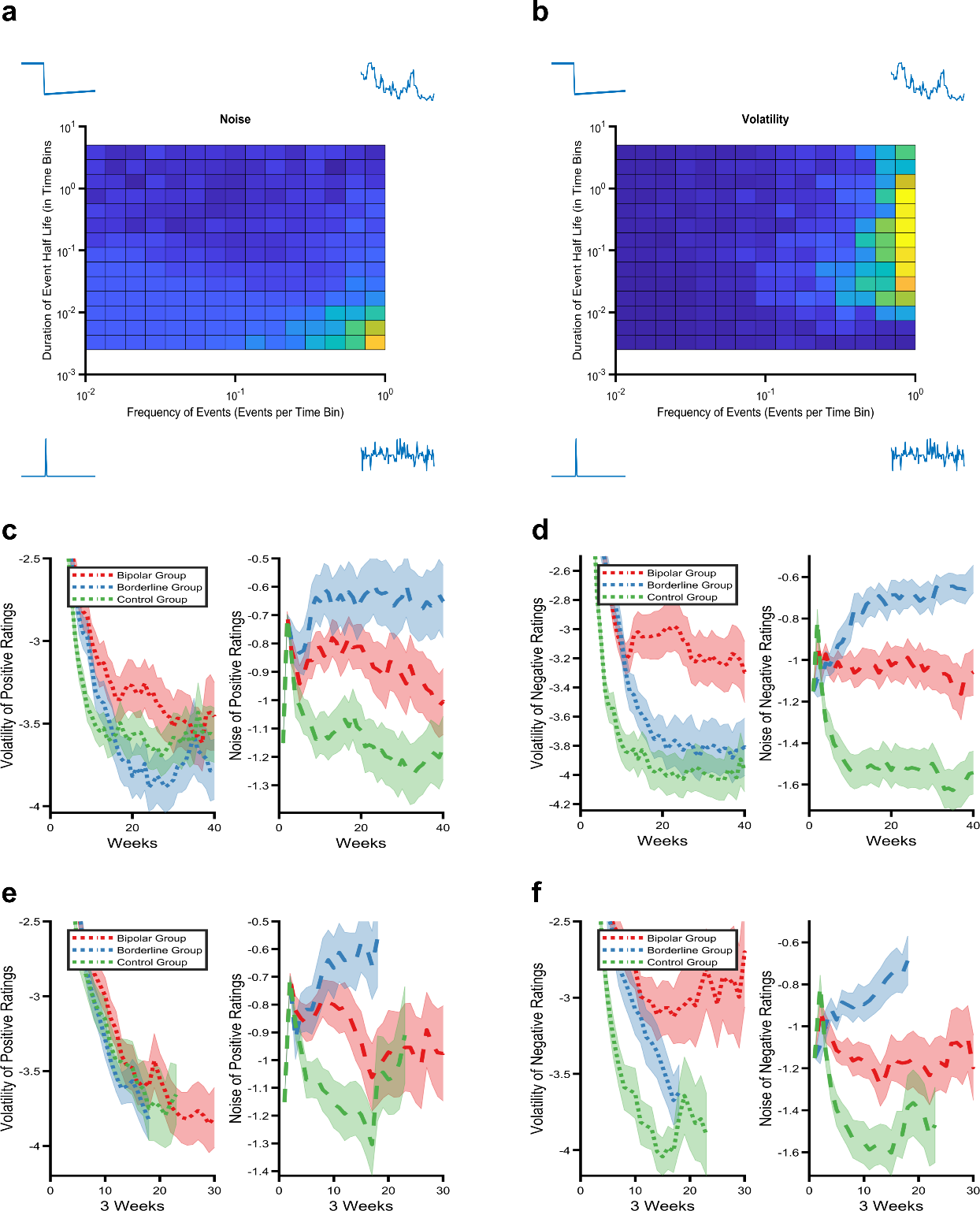

Figure S4: The relationship between the half life of affective perturbations and the filter derived metrics. Panels **a** and **b** illustrate the effect of the frequency of affective perturbations (x axis) and the duration of their effect (y axis) on estimates of volatility **a** and noise **b** from synthetic data. As can be seen, both volatility and noise increase as the frequency of affective perturbations increases, whereas the duration of the perturbations influences whether affective variability is attributed to volatility (for longer lasting perturbations) or noise (for shorter perturbations). Illustrative timecourses of the synthetic data are shown in the corner of each panel. Panels **c-f**. Group differences in the cause of affective variability in the cohort study are apparent when a lower sampling frequency of one (panels **c** and **d**) or three (panels **e** and f) weeks. The x-axis reports the number of time points (i.e. for the three week data time point 2 occurs three weeks after time point 1). Mean (SEM) of filter derived estimates are displayed as per the figures in the main paper, lines are censored when fewer than 10 participants are left in a group (i.e. participants remained in the study for variable lengths of time).

1.	T. E. J. Behrens, M. W. Woolrich, M. E. Walton, M. F. S. Rushworth, Learning the value of information in an uncertain world. *Nat. Neurosci.* **10**, 1214–1221 (2007).

2.	T. M. Liddell, J. K. Kruschke, Analyzing ordinal data with metric models: What could possibly go wrong? *Journal of Experimental Social Psychology* **79**, 328–348 (2018).

3.	E. Dejonckheere, *et al.*, Complex affect dynamics add limited information to the prediction of psychological well-being. *Nat Hum Behav* **3**, 478–491 (2019).
